# COVID-19 vaccination-induced antibody responses and waning by age and comorbidity status in a large population-based prospective cohort study

**DOI:** 10.1101/2023.10.24.23297192

**Authors:** CE Hoeve, AJ Huiberts, B de Gier, SP Andeweg, G den Hartog, HE de Melker, SJM Hahne, JHHM van de Wijgert, S van den Hof, MJ Knol

## Abstract

**Background:** Information on the magnitude and duration of antibody levels after COVID-19 vaccination in different groups may be useful for prioritizing of additional vaccinations.

**Methods:** Serum samples were collected every six months in a prospective cohort study among adults in the Netherlands. Geometric mean concentrations (GMCs) of antibodies against the receptor binding domain of the SARS-CoV-2 spike protein were calculated after the primary series, first, and second booster vaccinations. Effects of age (18-59 vs 60-85 years) and medical risk conditions on GMC 2-6 weeks and 21-25 weeks after each vaccination, and on waning during 3-25 weeks after each vaccination, were estimated by linear regression.

**Results:** We included 20,816, 16,820 and 5,879 samples collected after primary, first and second booster vaccination, respectively. GMCs at 2-6 and 21-25 weeks after primary series were lower in participants with older age or medical risk conditions. After the first booster, older age was associated with lower GMC at 2-6 weeks, higher GMC at 21-25 weeks, and slower waning. GMCs or waning after the first and second boosters (only 60-85) were not associated with medical risk conditions.

**Conclusions:** Since antibody differences by age and medical risk groups have become small with increasing number of doses, other factors such as disease severity rather than antibody levels are useful for prioritization of additional vaccinations.

## Introduction

COVID-19 vaccines provide protection against SARS-CoV-2 infection and against severe COVID-19 (1). The vaccines stimulate the immune system to produce protecting antibodies against the receptor binding domain of the SARS-CoV-2 S1 protein (S-antibodies). It has been shown that COVID-19 vaccines produce robust S-antibody responses after the primary series, and these responses are increased further following booster vaccinations. S-antibody concentrations have been reported to be higher for mRNA vaccines compared to viral vector vaccines and varying results were reported on the magnitude and/or duration of S-antibody responses by age and underlying medical conditions (2-7). Assessing the antibody response in vulnerable groups is valuable since SARS-CoV-2 S-antibody concentrations may be an indicator for protection against infection (5, 8, 9). Indeed, the Dutch Vaccine Study COVID-19 (VASCO) population-based cohort showed a dose-response relationship between S-antibody concentrations and protection against infection (10). After the roll-out of COVID-19 vaccination campaigns in the Netherlands from January 2021 onwards, waning of vaccine effectiveness against severe disease and of antibody concentrations has been reported (1). Booster campaigns have been implemented since November 2021 in an effort to prevent new waves of hospitalizations.

Booster doses have been reported to produce higher antibody concentrations than after the previous dose (5, 11). Age-dependent antibody responses have been reported after the primary series, with younger persons reaching higher levels (2, 5, 11). In the study by Wei et al, this effect was also observed after the first booster (5). However, differences between age groups have been reported to become smaller with increasing number of doses (2, 11). In the Netherlands, people with asplenia, diabetes, cardiovascular disease, immune disorder, cancer with current or no treatment, lung disease or asthma, hepatic disease, neurological disease, renal disease, organ or bonemarrow transplantation were prioritized for the primary series and booster vaccinations. Such medical risk groups are often prioritized for vaccination, but data on their response to vaccination are scarce.

The majority of studies describing antibody responses following COVID-19 vaccination are relatively small (<1000 participants), have a duration of follow up of only a few weeks and often focus on healthcare workers or patients with specific comorbidities. VASCO is a large population-based cohort study of approximately 45,000 participants, coordinated by the National Institute of Public Health and the Environment in the Netherlands. The study has been collecting longitudinal data on COVID-19 vaccinations, positive SARS-CoV-2 tests, and antibody concentrations since May 2023, 4 months after the start of the Dutch COVID-19 vaccination program. This allows evaluation of the humoral immune response following vaccination for multiple doses including the second booster dose while being able to differentiate between participants with and without prior infections. Participants over 60 years of age were oversampled, and the large sample size permits stratification by medical risk group. Therefore, this cohort can provide valuable information to support decision-making on the timing of booster vaccinations in the general population and by age and medical risk groups. This study investigates S-antibody concentrations shortly after and approximately 6 months after primary series, first, and second booster vaccinations, as well as the waning of S-antibody concentrations by vaccine product, having had a prior infection, age, and having at least one medical risk condition.

## METHODS

### Study design

VASCO is a prospective cohort study among ∼45,000 community-dwelling Dutch adults between 18-85 years (12). Participants were enrolled between 3 May 2021 and 15 December 2021 and are followed for five years. Data is collected through monthly questionnaires during the first year, and 3-monthly questionnaires in the following years, using the study-specific mobile phone application or website. The questionnaires include questions about demographics, medical conditions, medication use, COVID-19 vaccinations and positive SARS-CoV-2 tests. Participants are encouraged to get tested when they become symptomatic or have been exposed to SARS-CoV-2. Participants could access testing free-of-charge at a local public testing site until April 2022. After that date, the study team has been providing self-testing kits to the participants. Participants are asked to report positive SARS-CoV-2 tests (PCR or (self-administered) antigen test) and COVID-19 vaccinations using the study app. At enrollment, 6 months, 12 months, and 18 months follow-up, and one month following the primary vaccination series (for those who completed the primary vaccination series after enrollment), participants were asked to take a finger prick blood sample at home using a sampling kit received from the study team and mail it to the study laboratory. Participants were asked to report the sampling date in the study app. If the sampling date was missing, the date of sample receipt by the laboratory was used.

### Study population

For the current analysis, we used data collected between the study start in May 2021 and December 2022. Samples with missing data on age, medical risk group, vaccination status, and/or S-antibody blood concentrations were excluded. Five registered COVID-19 vaccines were used in the Netherlands during the study period: the mRNA vaccines Comirnaty (BNT162b2; BioNTech/Pfizer, Mainz, Germany/New York, United States (US)) and Spikevax (mRNA-1273, Moderna, Cambridge, US), the vector based vaccines Vaxzevria (ChAdOx1-S; AstraZeneca, Cambridge, United Kingdom) and Jcovden (Ad26.COV2-S (recombinant), Janssen-Cilag International NV, Beerse, Belgium), and a recombinant nanoparticle vaccine Nuvaxovid (NVX-CoV2373; Novavax CZ; Jevany, Czech republic). Participants could not choose which vaccine types they wanted as this was determined by the government based on safety/efficacy information and availability. Participants and their samples were excluded from the current analyses if more than 90 days had passed between the two primary series doses, or if the primary series was completed with a vaccine other than Comirnaty, Spikevax or Vaxzevria (due to the infrequent use of other vaccines). Furthermore, samples were excluded from participants who were unvaccinated, or whose primary series was incomplete, at the sampling date. Samples after booster vaccinations were excluded if the booster dose was a vaccine other than Comirnaty or Spikevax (these were the only booster vaccines administered in the governmental booster campaigns). Second booster analyses included samples of participants of 60 years or older only because younger persons without a medical condition were not eligible for the second booster. Samples taken after a third booster dose were excluded because we did not capture a sufficient number of those during our study period. Samples of participants who had a SARS-CoV-2 infection (as defined below) after vaccination and before the sampling date were excluded because the analyses focused on the effects of vaccinations. In case there were multiple samples from the same participant per dose, only the first sample per dose was included.

### Serum antibody measurements

Serum samples were analyzed with the Elecsys Anti-SARS-CoV-2 S and Anti-SARS-CoV-2 N assays on the Cobas e801 (Roche Diagnostics, Mannheim, Germany), which are electrochemiluminescence immunoassays measuring total immunoglobulin (Ig) levels against respectively the receptor binding domain (RBD) of the S1 protein (S-antibodies) and the nucleoprotein (N-antibodies) of SARS-COV-2. For S-antibody concentrations, the numeric results in U/mL of the Elecsys Anti-SARS-CoV 2 S assay are equivalent to binding antibody units (BAU/ml) and hence reported as such (Elecsys Anti-SARS-CoV 2 S method sheet 2022-02, V3.0). Samples with S-antibody concentrations higher than the upper limit of detection (up to 12,500 BAU/ml) were diluted 1:900, re-measured and quantified up to 225,000 BAU/ml. The lower detection limit was 20 BAU/ml. Measurements truncated at the lower or upper limit of detection were given the value of the limit of detection. Of the 43,515 samples, 65 were truncated at the lower limit and 817 were truncated at the upper limit. Seropositivity was defined as S-antibody concentrations of >20 BAU/ml. For N-antibody concentrations, the qualitative cut-off index was converted to numeric results in BAU/ml using batch-specific, linear calibration-lines obtained with a dilution range of the NIBSC 20/136 WHO standard or an internal pool of 125 N-antibody positive, anonymized patient sera calibrated against the WHO standard. The cut-off for N-positivity was set by converting cut-off index 1.0 to corresponding BAU/ml using these calibration lines.

### Infection and vaccination status

Infection and vaccination status were determined for each sample on the sampling date. For each sample a participant was classified as having had at least one prior infection preceding the sampling if 1) there was a self-reported positive SARS-CoV-2 test result at any time before sampling ; or 2) if the sample became seropositive for N-antibodies during the sampling interval or during a prior interval, or 3) if the N-antibody concentration increased at least four-fold during the sampling interval or during a prior interval. The date of infection was defined as the self-reported date of the positive SARS-CoV-2 test, or the mid-date between the current and previous sampling dates, respectively. When the first sample of a participant contained N-antibodies, but no positive test had been reported, the infection date was imputed as the mid-date between the baseline questionnaire and sample receipt. The infection date was used to determine whether the infection was before or after the last vaccination prior to the sampling date. Samples with an infection date after vaccination but before sampling were excluded.

Vaccination status was defined as primary series if the participant was vaccinated with only two primary doses; first booster if the participant was vaccinated with two primary doses plus one dose after 18 November 2021 (start of the first booster campaign in the Netherlands); second booster if the participant was vaccinated with two primary doses, one dose after 18 November 2021 and one dose after 26 February 2022 (start of the second booster campaign in the Netherlands) and before 19 September 2022 (start of the third (bivalent) booster campaign in the Netherlands). Vaccination data were based on self-reported vaccinations combined with data from the national vaccination register (COVID-vaccination Information and Monitoring System [CIMS]) as described previously (13).

### Medical risk group

Participants reporting at least one of the following conditions were included in the medical risk group: asplenia, diabetes, cardiovascular disease, immune disorder, cancer with current or no treatment, lung disease or asthma, hepatic disease, neurological disease, renal disease, and organ or bonemarrow transplantation. The following conditions were considered as immunocompromising conditions: immune disorder, cancer with current treatment, renal disease, and organ or bonemarrow transplantation. The presence of medical risk conditions was determined on the sampling date.

### Data-analysis

Characteristics of included participants were summarized by vaccination dose. To describe S-antibody concentrations over time since vaccination, log-transformed concentrations were modeled using a generalized additive model (GAM) for each dose and each vaccine product separately. To study the effect of vaccination only, participants with an infection at any time before vaccination were excluded from this analysis. Models were adjusted for age as a continuous variable interacting with time since vaccination in a tensor product with penalized splines (5 knots for age and 15 knots for time since vaccination) (14). The tensor products were allowed to vary by vaccine product, medical risk group, and sex. Predicted mean values with 95% CI were generated per day from which the day of peak response was determined and graphs for visual inspection were made. The predicted day of peak response was used for linear regression models on waning.

S-antibody seroconversion rates (from seronegative to seropositive) were calculated for each vaccine product and for participants with and without prior infection, using samples taken 14 to 42 days (2-6 weeks) following each vaccination. S-antibody responses 2-6 weeks (14-42 days) and 21-25 weeks (147-175 days) after vaccination were presented as geometric mean concentrations (GMC) by vaccine product, dose, prior infection, age group (18-59 years and 60-85 years) and medical risk group. Normality of log-transformed S-antibody concentrations was tested using a Shapiro-Wilk test. Linear regression on log-transformed S-antibody concentrations was done to test for differences between vaccine products stratified by dose and prior infection, adjusted for sex, age group and medical risk group, and to test for differences between medical risk and age groups stratified by vaccine product, dose, and prior infection, adjusted for sex. The resulting estimates were exponentiated to obtain GMC ratios with 95% CIs.

The effects of age group, medical risk group, and prior infection on waning were estimated by linear regression with interaction terms between time since vaccination and a combined age-medical risk group variable with four categories (18-59 years old without medical risk, 18-59 years old with medical risk, 60-85 years old without medical risk and 60-85 years old with medical risk), adjusted for sex and primary series vaccine product (the latter only in booster dose models). For this analysis, samples collected between the predicted S-antibody peak day from the GAM model and 175 days after vaccination were included. The models were stratified by dose and vaccine product. Samples with time since vaccination (in days) after the 95th percentile by age group and medical risk group were excluded, to avoid outliers. The effect of prior infection on waning was estimated by linear regression with an interaction term between time since vaccination and prior infection status, and adjusted for sex, age, medical risk group and primary series vaccine product (the latter only in booster dose models). Samples with time since vaccination (in days) after the 95th percentile, by infection status, were excluded, to avoid outliers. The resulting estimates were multiplied by 30 and exponentiated to obtain 30-day GMC ratios with 95% CIs.

All analyses were done using R version 4.2.2, using package mgcv.

## Ethical statement

The VASCO study protocol was approved by the not-for-profit independent Medical Ethics Committee of the *Stichting Beoordeling Ethiek Biomedisch Onderzoek* (BEBO), Assen, the Netherlands (NL76815.056.21) (12). Written informed consent was obtained from all participants, including consent for linking with the national registration of COVID-19 vaccination data (12). Further details on data management, privacy and ethics regarding the VASCO study are described by Huiberts et al (12).

## RESULTS

### Study population

A total of 43,515 finger prick samples collected from 29,732 participants were included of which 20,816 (48%) samples were taken following a primary series, 16,820 (39%) samples following a first booster, and 5,879 (14%) samples following a second booster (Table 1). There were slightly more samples from participants aged 60-85 years compared to 18-59 years for the primary series and first booster dose (Table 1). The analysis on the second booster dose was restricted to participants of 60 years and older, the main target group for this dose. The proportion of participants in the medical risk group was higher for those receiving the second booster (41%; 60-85 year olds only) than for those receiving the primary series and first booster (32% and 33%, respectively). The proportions of participants with medical risk conditions in the 18-59 years and 60-85 years groups were 23% and 39% for the primary series, and 25% and 38% for the first booster, respectively. Comirnaty was the most commonly used vaccine for the primary series, and Spikevax for the first and second boosters (Table 1). Vaccine products received by the participants varied by age and to some extent by medical risk group (supplementary table A). An infection was identified prior to the primary series vaccinations for 9% of samples and this increased with each dose to 35% of the samples taken after the second booster. The S-antibody GMCs increased with each vaccination dose from 1,390 BAU/ml (95%CI: 1,361;1,419) 2-6 weeks following the primary series to 22,975 BAU/ml (95%CI: 22,417;23,548) 2-6 weeks following the second booster.

**Table 1.** Description of participant characteristics by dose (n=29,732 participants). Participants may contribute multiple samples, up to a total of three samples, one after each vaccination dose.

|  | Primary series<br>(n=20,816 samples) | First booster<br>(n=16,820 samples) | Second booster<br>(n=5,879 samples) |
| --- | --- | --- | --- |
| **Age group** |  |  |  |
| 18-59 | 8,778 (42.2%) | 6,130 (36.4%) | N/A |
| 60-85 | 12,038 (57.8%) | 10,690 (63.6%) | 5,879 (100.0%) |
| **Sex** |  |  |  |
| Male | 7,763 (37.3%) | 6,074 (36.1%) | 2,561 (43.6%) |
| Female | 13,053 (62.7%) | 10,746 (63.9%) | 3,314 (56.4%) |
| **Medical risk group** | 6,714 (32.3%) | 5,597 (33.3%) | 2,399 (40.8%) |
| **Immune compromising condition\*** | 741 (3.6%) | 578 (3.4%) | 211 (3.6%) |
| **Vaccine product administered** |  |  |  |
| Comirnaty | 11,678 (56.1%) | 6,465 (38.4%) | 1,506 (25.6%) |
| Spikevax | 2,981 (14.3%) | 10,355 (61.6%) | 4,373 (74.4%) |
| Vaxzevria | 6,157 (29.6%) | N/A | N/A |
| **Infection at any time prior to** | 1,796 (8.6%) | 2,711 (16.1%) | 2,069 (35.2%) |

| vaccination |  |  |  |
| --- | --- | --- | --- |
| Median number of days between vaccine administration and sampling date [IQR] | 51 [34, 132] | 25 [12, 90] | 63 [40, 85] |
| GMC of S-antibodies in BAU/ml (95%CI) | 1,390 [1,361; 1,419] | 11,330 [11,079; 11,587] | 22,975 [22,417; 23,548] |

\* Immune disorder, cancer with current treatment, renal disease or organ/bonemarrow transplant

The S-antibody GMC increased in the first three weeks after vaccination and then showed a steady decline for all vaccine products, age groups, medical risk groups and doses (Figure 1). The predicted day on which the peak GMC was reached was between 15 and 22 days post-vaccination for all vaccines and doses. The 22^nd^ day was used as the starting point for the models on waning.

**Figure 1.**
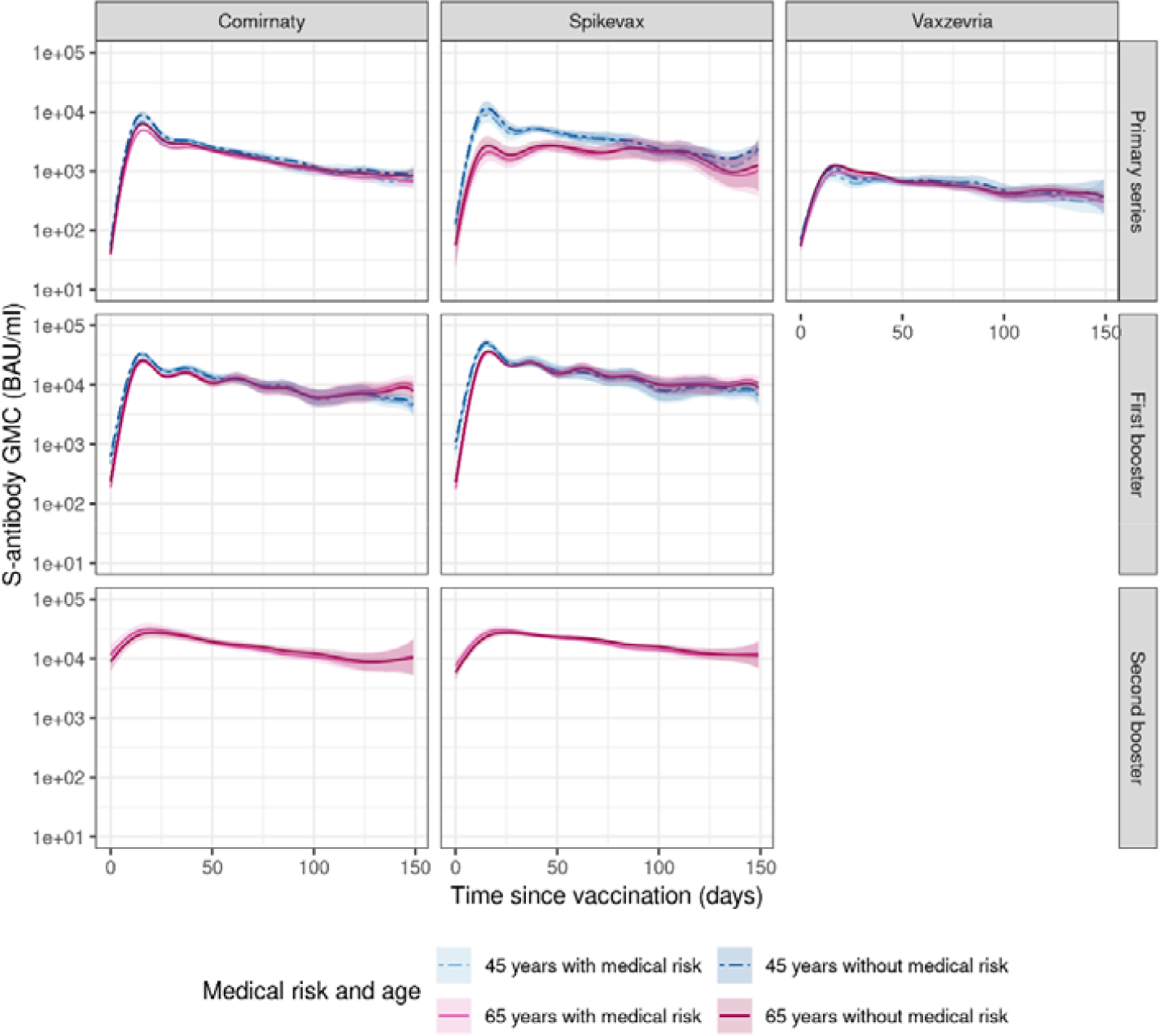
Predicted S-antibody GMC over time by vaccine product, dose, age, and medical risk group in individuals without prior infection. Plotted data are based on a generalized additive model, colored lines represent predicted S-antibody GMC over time for women of age 45 and 65 and with and without medical risk conditions accompanied by 95% confidence intervals. Ages 45 and 65 were the median ages in the 18-59 and 60-85 year age groups, respectively.

### Seropositivity and S-antibody response shortly after vaccination

Over 98% of samples of participants uninfected prior to vaccination, taken between 2-6 weeks following the primary series, were seropositive for S-antibodies (ranging from 98.0% for Vaxzevria to 99.4% for Comirnaty, supplementary table B). Almost all samples (99.9%), were seropositive after the first booster and all samples (100.0%) after the second booster. Samples from participants who had an infection prior to vaccination were almost always (99.2%-100.0%) seropositive 2-6 weeks after the primary series, and all were seropositive 2-6 weeks after the first booster.

In general, S-antibody GMCs were higher in participants with a prior infection, vaccinated with mRNA vaccines, under 60 years of age, and not in the medical risk group. Differences became smaller after the booster doses.

More specifically, linear regression showed that the differences in GMC 2-6 weeks after vaccination between vaccine products were all significant, except after the second booster (supplementary table C), both for samples with and without infection prior to vaccination. In samples without infection prior to vaccination, GMC 2-6 weeks following primary series was highest for Spikevax (GMC [95%CI]: 4,077 [3,612-4,602]) followed by Comirnaty (GMC [95%CI]: 3,038 [2,943-3,136]) and Vaxzevria (GMC [95%CI]: 756 [712-804]). GMC 2-6 weeks following a Spikevax booster (GMC+95%CI: 24,503 [23,852-25,172]) was significantly higher than the GMC 2-6 weeks following a Comirnaty booster (GMC [95%CI]: 18,666 [18,001 -19,356]). The difference in GMC between Comirnaty and Spikevax at 2-6 weeks following a second booster was smaller and not significantly different, owing mostly to a relative increase of GMC in the Comirnaty group (GMC [95%CI]: 23,889 [20,944 -27,248] vs. GMC [95%CI]: 25,459 [23,664 -27,391] for Spikevax). A similar pattern was observed among samples with an infection prior to vaccination, although GMCs were generally higher: the Spikevax GMC was also significantly higher than the Comirnaty and Vaxzevria GMCs following primary series and higher than the Comirnaty GMC following first booster in individuals with prior infection, but was not different from the Comirnaty GMC after the second booster.

After stratifying by vaccine product some differences were observed by age and medical risk group (figure 2, supplementary table D). For those without prior infection receiving a primary series with Comirnaty or Spikevax, S-antibody GMCs were significantly lower in the 60-85 years age group and in those in the medical risk group. For example, for Comirnaty the GMC ratio for participants of 60-85 years compared to 18-59 years was 0.833 (95%CI: 0.769-0.902) and the GMC ratio for participants with medical risk conditions compared to those without was 0.844 (95%CI: 0.746-0.953). No differences were seen between the age and medical risk groups receiving a Vaxzevria primary series. After the first booster, regardless of vaccine product, S-antibody GMCs were significantly lower in the 60-85 years age group (e.g. the GMC ratio for Comirnaty was 0.787 [95%CI: 0.722-0.858]) but not in the medical risk group. After the second booster, there was no difference between 60-85 year old participants with or without medical risk conditions. Among those with prior infection, S-antibody GMCs were significantly higher in the older age group if they were receiving Vaxzevria (GMC ratio: 2.206 [95%CI: 1.054-4.609]) and significantly lower in the older age group if they were receiving Spikevax as primary vaccination (GMC ratio: 0.084 [95%CI: 0.029-0.243]), however numbers were low in the Spikevax group with prior infections. In those with prior infection before a first booster, the age effect that was seen in those without prior infection was not observed. For those with a Comirnaty first booster the S-antibody GMC was significantly higher in participants of 60-85 years with medical risk conditions (GMC ratio: 1.576 [95%CI: 1.260-1.972]). No differences were observed for the medical risk group after the second booster for those with a prior infection.

**Figure 2.**
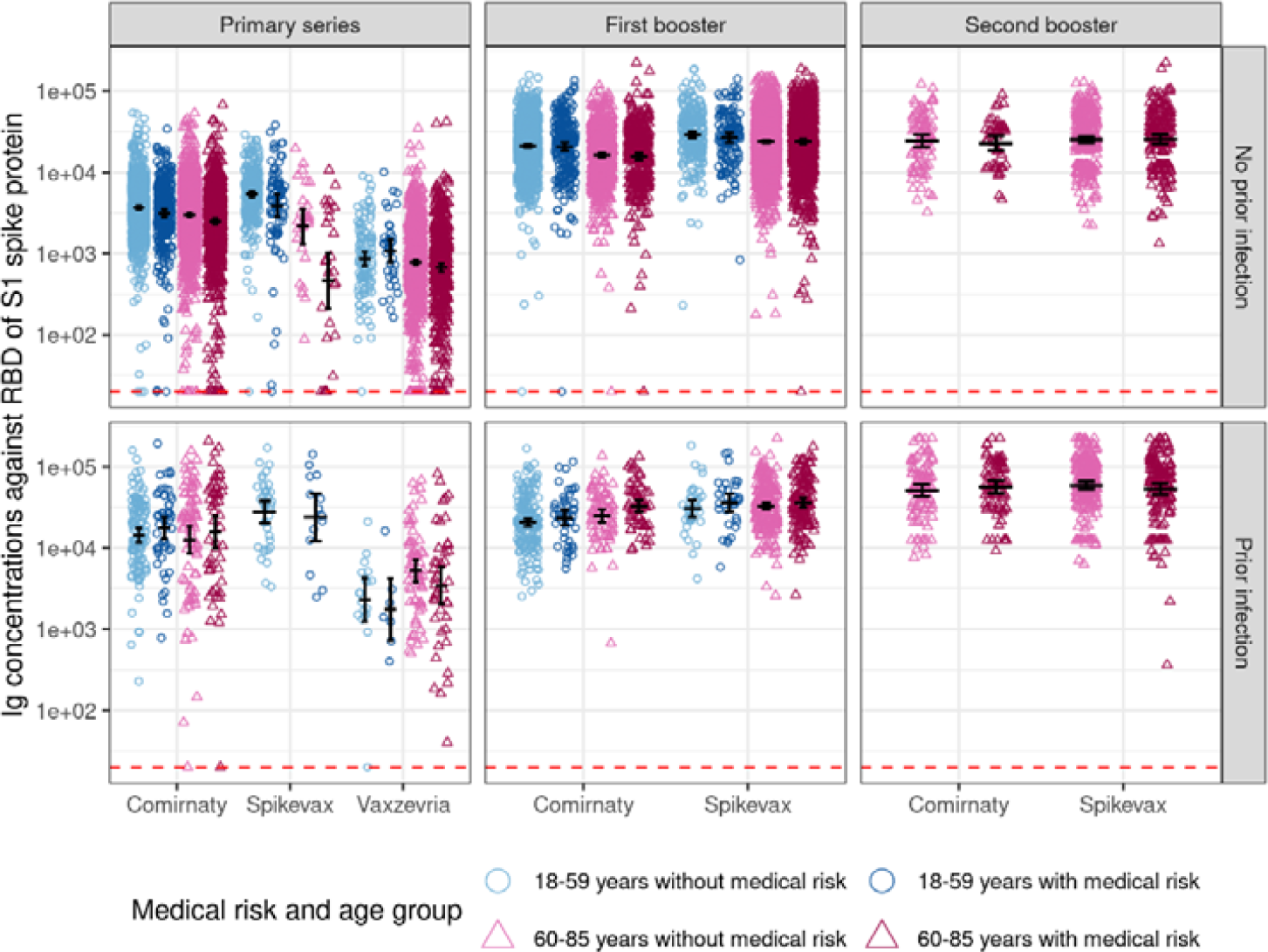
S-antibody levels 2-6 weeks following vaccination by vaccine product, dose, having had at least one prior infection, age group and medical risk group. Medical risk group is defined as asplenia, diabetes, cardiovascular disease, immunedisorder, cancer with current or no treatment, lung disease or asthma, hepatic disease, neurological disease, renal disease, organ or bonemarrow transplant. Black lines represent GMC + 95% CI, red-dotted lines represent S-antibody concentrations at which participants are considered seropositive. Groups with <5 observations are not shown.

### Waning of S-antibody response

We estimated the effects of age group and medical risk group on waning and included only samples without infection prior to vaccination (see supplementary table E for all estimates). Based on the model estimates, we plotted the waning until 175 days after vaccination by vaccine and dose for female participants in the 18-59 years and 60-85 years age groups, with and without medical risk conditions (figure 3). Since waning was modelled independent of sex and female participants were more frequent, female data is shown in the figures.

**Figure 3.**
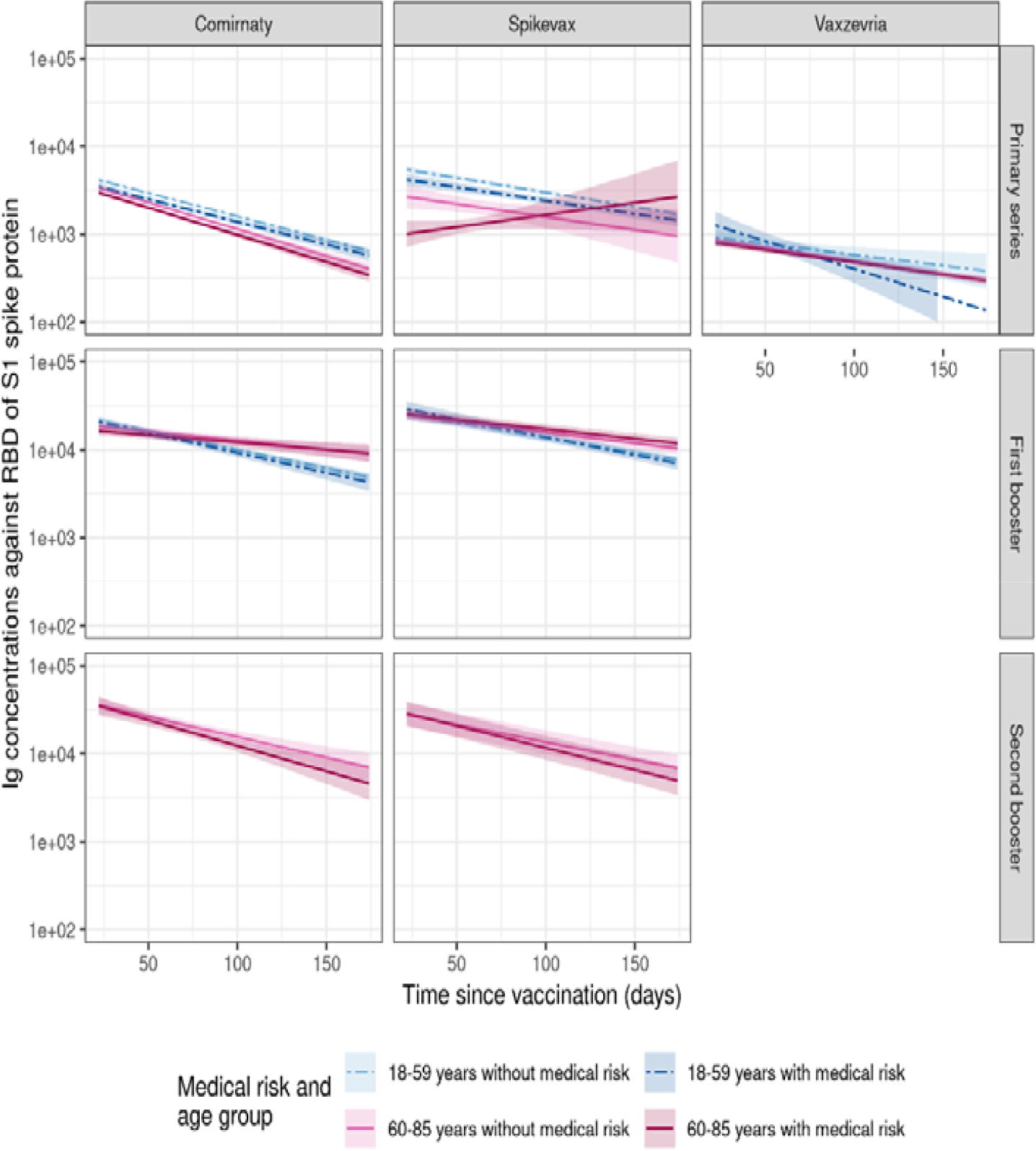
Estimated effect of age and medical risk conditions on waning. Figures present data for female participants in the age groups 18-59 years and 60-85 years, with and without medical risk conditions. Waning is presented by vaccine product and dose. Colored lines represent estimated mean Ig with 95% CI. The model for the second booster was only based on data from participants over 60 years.

Waning after primary series was significantly faster in the medical risk group after vaccination with Vaxzevria (0.767 95%CI: 0.590-0.996). In addition, we observed that waning after primary series was significantly faster in the 60-85 year age group following vaccination with Comirnaty (0.951 95%CI: 0.913-0.991), but slower in the 60-85 year age group if they also had medical risk conditions following vaccination with Spikevax (1.524 95%CI: 1.191-1.950). After the first booster, waning was significantly slower in the 60-85 year age group (Comirnaty 1.161 95% CI: 1.101-1.225; Spikevax 1.064 95%CI: 1.019-1.111) and there was no effect of medical risk conditions on waning, regardless of vaccine product. Following the second booster (only in the 60-85 year age group) no effect of medical risk condition on waning was observed.

Waning patterns were slightly different when using models with a smaller medical risk group definition (with only immune compromising conditions, see supplementary table F). Waning following the primary series was only faster among Comirnaty-vaccinated participants without immune compromising conditions in the 60-85 year age group. Similarly to the previous model, waning following the first booster was slower in participants in the 60-85 year age group (Comirnaty 1.177 95%CI: 1.127-1.229; Spikevax 1.091 95%CI: 1.053-1.131). However, we also observed faster waning among the Comirnaty-vaccinated participants with immune compromising conditions in the 18-59 year age group (0.842 95%CI: 0.728-0.973). Waning following the second booster was faster among Spikevax receiving participants with immune compromising conditions (0.825 95%CI: 0.693-0.982).

Estimates of the effect of prior infection on waning are presented in the supplementary table G. Based on the model estimates, we plotted the waning until day 175 for female participants in the 60-85 year age group and without medical risk conditions, with and without infection prior to vaccination (figure 4). The effect of prior infection was not consistent across different doses or vaccines. Infection prior to vaccination was associated with faster waning following the Spikevax primary series (0.836, 95%CI: 0.781-0.896) and slower waning following the Vaxzevria primary series (1.152, 95%CI: 1.079-1.231) and Comirnaty first booster (1.130, 95%CI: 1.082-1.179). For other doses no significant effect of prior infection on waning was observed. After 175 days S-antibody levels in participants with prior infection were still higher than in those without prior infection.

**Figure 4.**
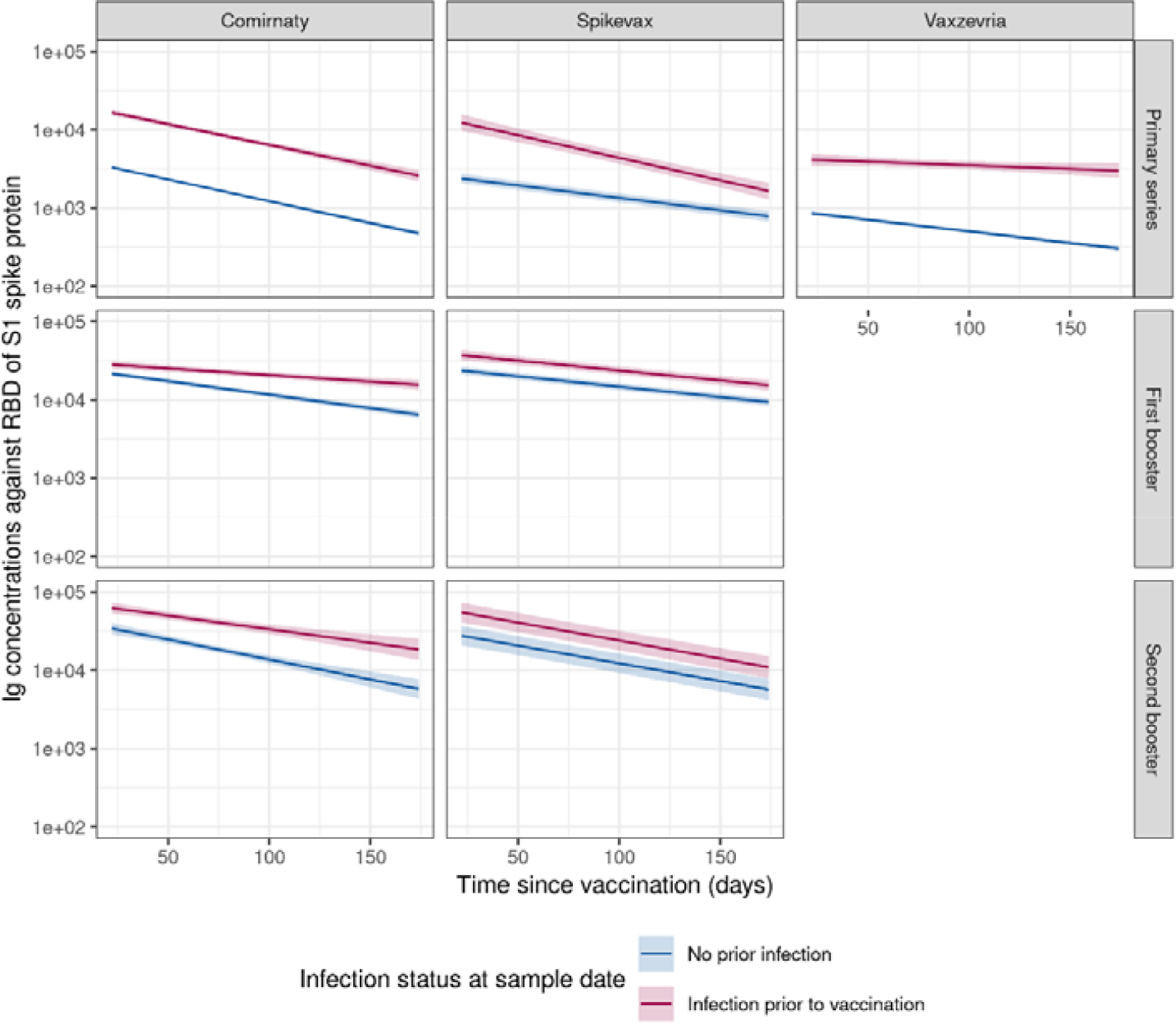
Estimated effect of prior infection on waning for female participants in the 60-85 year age group and without medical risk group indication. Waning is presented by vaccine product and dose. Colored lines represent estimated mean Ig with 95% CI. The model for the second booster was only based on data from participants over 60.

### S-antibody response 21-25 weeks after vaccination

In general, after 21-25 weeks following primary vaccination, antibody levels were lower in participants over 60 years compared to those under 60 years. However, 21-25 weeks after the first booster dose antibody levels were higher in participants over 60 years. Medical risk conditions did not significantly affect antibody levels after 21-25 weeks.

More specifically, after 21-25 weeks GMC was much reduced compared to the GMC after 2-6 weeks for all vaccine products and doses (figure 5). Among those without infection before vaccination, GMC at 21-25 weeks following primary vaccination with Comirnaty or Spikevax was lower in the older age group. For Vaxzevria there were no differences in GMC for the primary vaccination between age groups or those with and without medical risk conditions. After the first booster GMC was significantly higher in the older age group regardless of vaccine product (Comirnaty or Spikevax). There were no significant differences in GMC between groups after the second booster. For those with an infection before vaccination there were no differences between groups after the primary vaccination, regardless of vaccine. However, after the first booster GMC at 21-25 weeks was significantly higher in the older age group for both Spikevax and Comirnaty. There were no significant differences in GMC between groups after the second booster.

**Figure 5.**
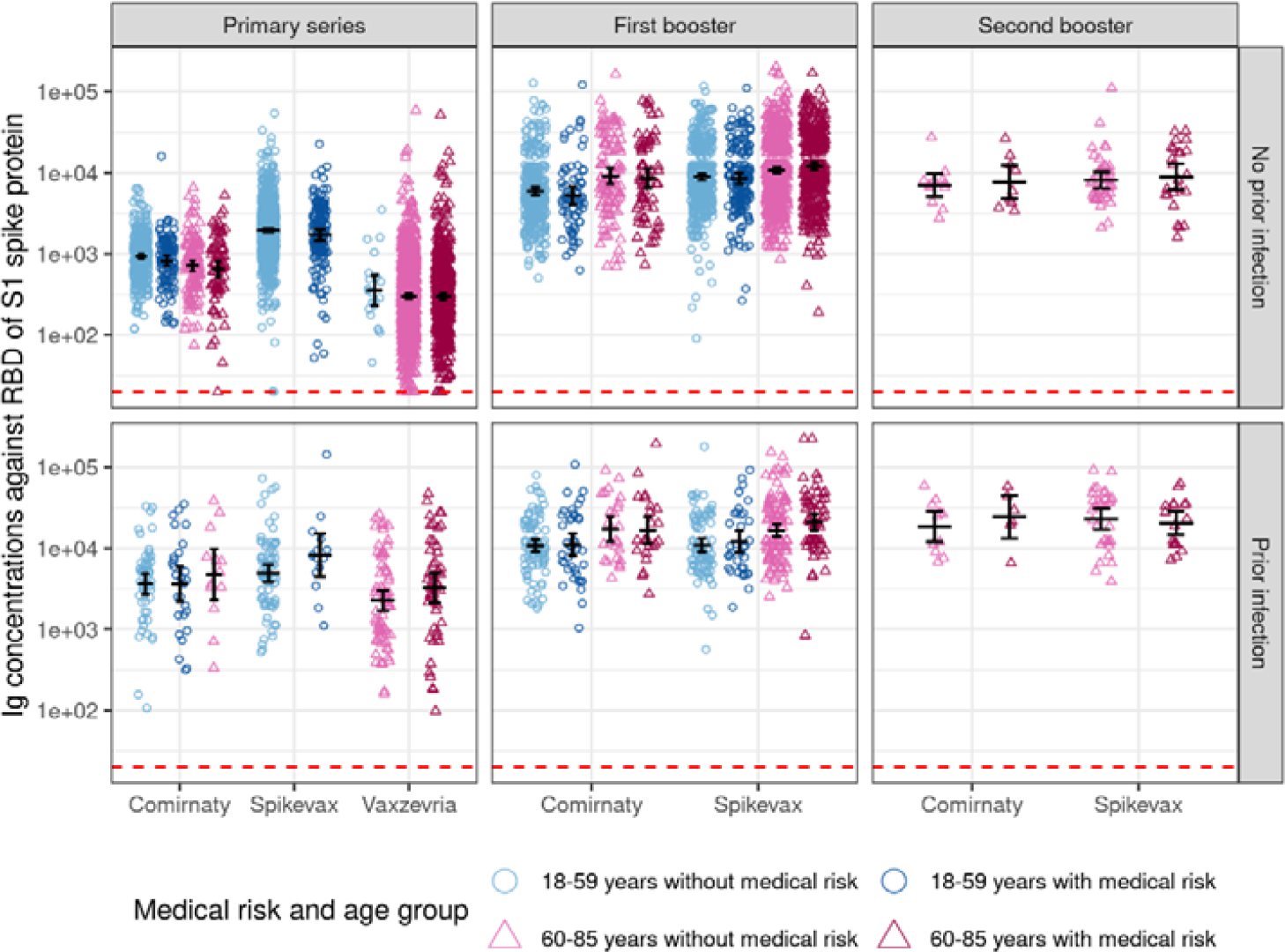
S-antibody levels 21-25 weeks following vaccination by vaccine product, dose, prior infection status, age group and medical risk group. Medical risk group is defined as asplenia, diabetes, cardiovascular disease, immunedisorder, cancer with current or no treatment, lung disease or asthma, hepatic disease, neurological disease, renal disease, organ or bonemarrow transplant. Black lines represent GMC + 95% CI, red-dotted lines represent S-antibody concentrations at which participants are considered seropositive. Groups with <5 observations are not shown.

## DISCUSSION

We used data from a large prospective cohort study to study determinants of S-antibody response and waning after primary and booster vaccinations. We found that after primary vaccination S-antibody levels at 2-6 weeks following primary vaccination with Spikevax or Comirnaty are respectively 60% and 17% lower in 60-85 year olds than in 18-59 year olds, 16-28% lower in participants with a medical condition, ∼5 times higher in participants with a prior infection and higher for the mRNA vaccines compared to Vaxzevria. Additionally, we observed slower waning of S-antibody levels in the older than younger age group following primary series with Comirnaty. We observed faster waning of S-antibody levels for participants with than those without medical risk conditions after the primary series with Vaxzevria. After 21-25 weeks since primary vaccination S-antibody levels were 23-52% lower in the 60-85 year age group than in the 18-59 year age group. The differences between vaccines and medical risk groups became smaller after the first booster, but S-antibody levels were still lower in the older age group. Additionally, we observed slower waning of S-antibody levels in the older than younger age group following first booster doses. As a result, S-antibody levels at 21-25 weeks after the first booster were 1.2-1.5 times higher in the 60-85 year age group compared to the younger group, in contrast to the lower levels in the elder group shortly after primary vaccination. Shortly following the second booster dose no differences were observed in S-antibody response between vaccines or medical risk groups for participants over 60 years (i.e. those targeted for the second booster dose). Waning after the second booster dose was not different in those with medical risk conditions compared to those without and after 21-25 weeks S-antibody levels were still similar in those with medical risk conditions compared to those without.

In line with our results, lower S-antibody levels shortly after vaccination among those of higher age or with medical conditions have been reported for the primary series (2-5, 15, 16). Also, Sanada *et al*. reported that the effect of age was no longer present following the first booster dose (11). We observed that S-antibody levels were still lower in the older age group shortly after the first booster dose. Surprisingly, waning was also slower for this group. This resulted in a lasting immune response in the older age group as we observed that 21-25 weeks since the first booster dose they actually had higher S-antibody levels compared to those in the younger age group. Unfortunately we could not confirm this effect for the second booster dose since the second booster campaign was only implemented in the older age group. These results are encouraging as they suggest a long lasting immune response in the age group most affected by severe disease from SARS-CoV-2 infections. We observed significantly lower S-antibody levels shortly following primary vaccination for participants with a medical risk condition. However, S-antibody levels at 21-25 weeks for those with medical risk or immune compromising conditions were not significantly different from those without following primary and also not for first, or second booster vaccination. Therefore, although the waning for this group appears to be a little faster shortly after (primary) vaccination, this does not seem to directly impact the long term antibody levels.

We earlier showed a dose-response relationship between S-antibody GMC and protection against infection (10). COVID-19 vaccines show waning vaccine effectiveness over time, as was previously also observed in the VASCO study (10), and in line with the observed waning of S-antibodies in this study. We also reported lower vaccine effectiveness among participants with medical risk conditions, but it was not obvious that the difference in VE between those with and without medical risks changed over time (10). This is in line with our finding that S-antibodies of those with and without medical risk do not wane at a different pace. A study by Wei *et al*. looked at an antibody threshold to relate to protection. They estimated that antibodies remained high enough for 60-100 days to give a 67% protection against infection, depending on age and vaccine product (5). Information on peak response levels and subsequent waning of immune response in risk groups could thus support decision making on the timing of booster vaccinations. In this context, since antibodies are not the only factor relevant for protection against SARS-CoV-2 infection (5, 8, 9, 17, 18), not only the level of protection but also the estimated severity of acquired infection in specific groups (e.g. >70 years or medical risk conditions) and other relevant factors in the protection against SARS-CoV-2 infection (e.g. cellular immunity) (5, 10, 13) should be considered.

After waning of S-antibody levels, further booster doses increase S-antibody levels again. We observed a marked increase in GMC shortly following the first booster compared to the primary series and a further increase after the second booster dose in line with previous findings (5, 11, 19-22). However, the increase in GMC shortly following the second booster compared to the first booster was much smaller. This suggests that additional doses may not lead to a much further increase of the peak antibody response. However, it should be noted that the time between boosters in our study period was relatively short (around 6 months). Perhaps, if time between boosters increases, peak response could be affected differently. Vaccine product could also potentially affect the peak levels that are reached. Although almost all participants seroconverted within 3 months following the primary series (regardless of vaccine product), we observed large differences between vaccines in GMC at 2-6 weeks following the primary series, with the highest response observed for Spikevax and the lowest for Vaxzevria, as has been reported in other studies (2-4, 6, 7, 23). A role for the different mechanisms of mRNA versus vector based vaccines has been hypothesized. After the first and second booster doses, Spikevax peak response remained higher than Comirnaty, although the difference became smaller. The decrease in difference of antibody response between Spikevax and Comirnaty following the booster dose has been described before (5). We show that this difference further narrows following the second booster. It has been hypothesized that this can partly be explained by the higher dose of Spikevax vaccines, which was 100 µg for the primary series doses and 50 µg for the booster doses versus 30 µg for Comirnaty (5). In an earlier study we showed that the vaccine effectiveness for Vaxzevria is lower than for Spikevax and Comirnaty (24), in line with the lower antibodies we found for Vaxzevria.

Prior infection is known to induce higher peak responses following subsequent vaccination (3, 5, 23) and was reported to be associated with slower waning of antibodies after primary series and first booster (5, 23). We showed earlier that hybrid immunity (i.e. a prior infection and vaccination) presents with higher protection against infection than only vaccination, but that waning of S-antibodies was faster than waning of protection (10). Our data also show higher peak antibody levels among participants with infection prior to vaccination, however, we found mixed results concerning waning in this group of participants. Following primary series vaccination our data suggests slower waning of antibodies for infected participants vaccinated with Vaxzevria or Comirnaty (although the latter was not significant), but faster waning for infected participants who were vaccinated with Spikevax compared to participants without infection prior to vaccination. Following the first booster, waning was only slower for participants who were infected prior to receiving Comirnaty. After the second booster, prior infection was not associated with waning. Based on our data we could not conclude whether the duration of S-antibody response is longer or shorter in those with an infection prior to vaccination. Regardless of waning, we also observed that GMC at 21-25 weeks after vaccination were still higher in the group with a prior infection, compared to those without.

The strength of our study lies in the large sample size of our prospective cohort with detailed information on participant characteristics, such as underlying conditions. Our study oversampled participants over 60 years old and is sufficiently large for stratification by medical risk groups allowing for in-depth analysis of this group that has a higher risk of severe disease from SARS-CoV-2 infection. A limitation of our study is the exclusion of people in care homes, whereas this might be the population most in need for repeated booster vaccination. To evaluate the antibody levels over time after vaccination, samples from different individuals were used rather than multiple samples from the same individual. Especially in smaller subgroups this may have led to spurious results. A total of 13% of the infections before vaccination were identified using only serology data, and date of infection was imputed leading to potential misclassification in identifying whether infections were before or after vaccination. However, using serology data we were also able to identify more infections than when using only reported testing data. We have potentially missed some infections (especially those before study enrollment) due to waning of N-antibodies over time and suboptimal sensitivity of the assay. In this study we included only participants with a history of COVID-19 vaccination. Vaccination data is self-reported and compared with the COVID-vaccination Information and Monitoring System (CIMS), in order to complete potentially missing vaccination data. In case no informed consent for data collection was given to CIMS or for coupling of study data with CIMS, comparison was not possible. However, this concerned only a small amount of participants. Medical risk group is defined using self-reported conditions which are used for inviting citizens for influenza vaccination. This concerns a heterogeneous group including conditions such as diabetes, cardiovascular disorders and immune compromising conditions. The latter group, for which we expected deviating patterns in S-antibody response, was therefore analyzed separately.

## Conclusion

Our data show long-lasting S-antibody levels in younger and older participants and for those in the medical risk group. Since the differences in S-antibody levels by age and medical risk groups have become small with increasing number of doses, other factors such as disease severity may become more relevant factors to use for prioritization of vaccination.

## Data Availability

All data produced in the present study are available in aggregated and anonymized form upon reasonable request to the authors.

