## Supplemental files for "COVID-19 vaccination-induced antibody responses and waning by age and comorbidity status in a large population-based prospective cohort study"

### SUPPLEMENTS

**Table A. Distribution of administered vaccines across medical risk group and age groups**

| Dose | Age group | Medical risk group | Comirnaty | Spikevax | Vaxzevria |
|---|---|---|---|---|---|
| Primary series | 18-59 | No | 4100 (60%) | 2250 (33%) | 428 (6%) |
| | 18-59 | Yes | 1319 (66%) | 508 (25%) | 173 (9%) |
| | 60-85 | No | 3665 (50%) | 100 (1%) | 3559 (49%) |
| | 60-85 | Yes | 2594 (55%) | 123 (3%) | 1997 (42%) |
| First booster | 18-59 | No | 3092 (67%) | 1526 (33%) | |
| | 18-59 | Yes | 842 (56%) | 670 (44%) | |
| | 60-85 | No | 1572 (24%) | 5033 (76%) | |
| | 60-85 | Yes | 959 (23%) | 3126 (77%) | |
| Second booster | 60-85 | No | 904 (26%) | 2576 (74%) | |
| | 60-85 | Yes | 602 (25%) | 1797 (75%) | |

**Table B. Rates of samples seropositive for S-antibodies by vaccine product, dose and prior infection based on finger prick samples taken 2-6 weeks following a vaccination.**

| Vaccine | 2-6 weeks since primary series | 2-6 weeks since booster 1 | 2-6 weeks since booster 2 |
|---|---|---|---|
| **No prior infection** | | | |
| Comirnaty | 3534/3554 (99.4%) | 2114/2118 (99.8%) | 126/126 (100%) |
| Spikevax | 402/409 (98.3%) | 3271/3272 (100%) | 455/455 (100%) |
| Vaxzevria | 1446/1475 (98%) | N/A | N/A |
| **Infection prior to vaccination** | | | |
| Comirnaty | 290/292 (99.3%) | 356/356 (100%) | 168/168 (100%) |
| Spikevax | 59/59 (100%) | 390/390 (100%) | 363/363 (100%) |
| Vaxzevria | 129/130 (99.2%) | N/A | N/A |

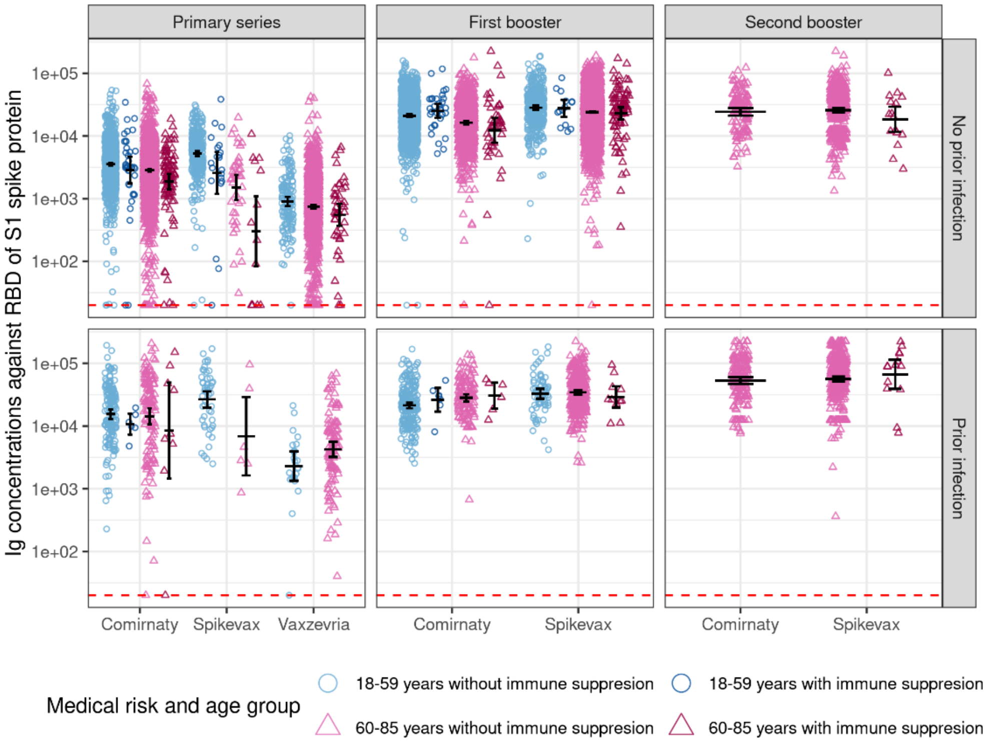


**Figure A. S-antibody levels 2-6 weeks following vaccination by vaccine product, dose, prior infection status, age group and immune compromising condition.** Immune compromising conditions are defined as immune disorder, cancer with current treatment, renal disease, organ or bonemarrow transplant. Black lines represent GMC + 95% CI, red-dotted lines represent S-antibody concentrations at which participants are considered seropositive. Groups with <5 observations are not shown.

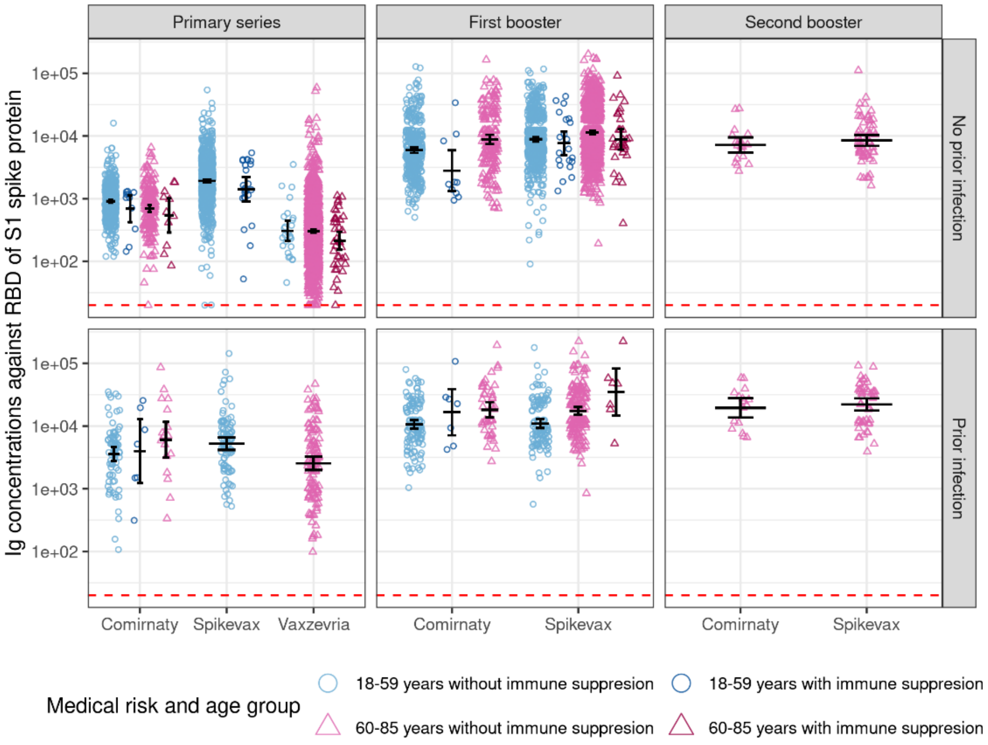


**Figure B. S-antibody levels 21-25 weeks following vaccination by vaccine product, dose, prior infection status, age group and immune compromising condition.** Immune compromising conditions are defined as immune disorder, cancer with current treatment, renal disease, organ or bonemarrow transplant. Black lines represent GMC + 95% CI, red-dotted lines represent S-antibody concentrations at which participants are considered seropositive. Groups with <5 observations are not shown.

**Table C. Effect of vaccine products on antibody levels at 2-6 weeks following vaccination.** Models are stratified by prior infection and dose and adjusted for sex, age group and medical risk group.

| Prior infection | Dose | Vaccine product | GMC ratio at 2-6 weeks | 95% CI | n | GMC ratio at 21-25 weeks | 95% CI | n |
|---|---|---|---|---|---|---|---|---|
| No | Primary series | Comirnaty | Ref | | 3554 | Ref | | 657 |
| No | Primary series | Spikevax | 1,148 | 1,026 - 1,284 | 409 | 2,094 | 1,891 - 2,316 | 1052 |
| No | Primary series | Vaxzevria | 0,264 | 0,247 - 0,282 | 1475 | 0,429 | 0,376 - 0,489 | 1602 |
| No | First booster | Comirnaty | Ref | | 2118 | Ref | | 543 |
| No | First booster | Spikevax | 1,473 | 1,401 - 1,548 | 3272 | 1,415 | 1,276 - 1,570 | 1470 |
| No | Second booster | Comirnaty | Ref | | 126 | Ref | | 21 |
| No | Second booster | Spikevax | 1,065 | 0,911 - 1,244 | 455 | 1,157 | 0,791 - 1,694 | 64 |
| Yes | Primary series | Comirnaty | Ref | | 292 | Ref | | 101 |
| Yes | Primary series | Spikevax | 1,603 | 1,076 - 2,389 | 59 | 1,451 | 0,991 - 2,121 | 88 |
| Yes | Primary series | Vaxzevria | 0,264 | 0,194 - 0,358 | 130 | 0,467 | 0,263 - 0,828 | 123 |
| Yes | First booster | Comirnaty | Ref | | 356 | Ref | | 176 |
| Yes | First booster | Spikevax | 1,311 | 1,161 - 1,480 | 390 | 1,053 | 0,881 - 1,261 | 276 |
| Yes | Second booster | Comirnaty | Ref | | 168 | Ref | | 17 |
| Yes | Second booster | Spikevax | 1,050 | 0,897 - 1,230 | 363 | 1,080 | 0,700 - 1,664 | 48 |

**Table D. Effect of age and medical risk group on antibody levels at 2-6 weeks and 21-25 weeks following vaccination.** Models are stratified by vaccine product, prior infection and dose and adjusted for sex.

| Prior infection | Dose | Vaccine product | Age and medical risk group | GMC ratio At 2-6 weeks | 95% CI | n | GMC ratio At 21-25 weeks | 95% CI | n |
|---|---|---|---|---|---|---|---|---|---|
| No | Primary series | Comirnaty | 18-59 years without medical risks | Ref | | 955 | Ref | | 359 |
| No | Primary series | Comirnaty | 18-59 years with medical risks | 0,844 | 0,746 - 0,953 | 310 | 0,883 | 0,751 - 1,039 | 105 |
| No | Primary series | Comirnaty | 60-85 years without medical risks | 0,833 | 0,769 - 0,902 | 1355 | 0,767 | 0,658 - 0,896 | 120 |
| No | Primary series | Comirnaty | 60-85 years with medical risks | 0,710 | 0,650 - 0,775 | 934 | 0,691 | 0,573 - 0,834 | 73 |
| No | Primary series | Spikevax | 18-59 years without medical risks | Ref | | 280 | Ref | | 906 |
| No | Primary series | Spikevax | 18-59 years with medical risks | 0,720 | 0,547 - 0,948 | 76 | 0,889 | 0,768 - 1,027 | 140 |
| No | Primary series | Spikevax | 60-85 years without medical risks | 0,398 | 0,257 - 0,616 | 26 | 0,476 | 0,232 - 0,976 | 5 |
| No | Primary series | Spikevax | 60-85 years with medical risks | 0,083 | 0,054 - 0,128 | 27 | 0,328 | 0,066 - 1,624 | 1 |
| No | Primary series | Vaxzevria | 18-59 years without medical risks | Ref | | 110 | Ref | | 21 |
| No | Primary series | Vaxzevria | 18-59 years with medical risks | 1,242 | 0,795 - 1,941 | 36 | 0,476 | 0,166 - 1,365 | 5 |
| No | Primary series | Vaxzevria | 60-85 years without medical risks | 0,919 | 0,726 - 1,165 | 843 | 0,875 | 0,548 - 1,395 | 1011 |
| No | Primary series | Vaxzevria | 60-85 years with medical risks | 0,796 | 0,622 - 1,020 | 486 | 0,869 | 0,543 - 1,394 | 565 |
| No | First booster | Comirnaty | 18-59 years without medical risks | Ref | | 918 | Ref | | 303 |
| No | First booster | Comirnaty | 18-59 years with medical risks | 0,989 | 0,877 - 1,116 | 235 | 0,881 | 0,669 - 1,161 | 67 |
| No | First booster | Comirnaty | 60-85 years without medical risks | 0,787 | 0,722 - 0,858 | 627 | 1,531 | 1,213 - 1,933 | 103 |
| No | First booster | Comirnaty | 60-85 years with medical risks | 0,770 | 0,692 - 0,857 | 338 | 1,439 | 1,095 - 1,889 | 70 |
| No | First booster | Spikevax | 18-59 years without medical risks | Ref | | 254 | Ref | | 395 |
| No | First booster | Spikevax | 18-59 years with medical risks | 0,917 | 0,776 - 1,083 | 127 | 0,970 | 0,808 - 1,166 | 159 |
| No | First booster | Spikevax | 60-85 years without medical risks | 0,853 | 0,770 - 0,945 | 1811 | 1,196 | 1,051 - 1,359 | 548 |
| No | First booster | Spikevax | 60-85 years with medical risks | 0,859 | 0,772 - 0,956 | 1080 | 1,335 | 1,158 - 1,537 | 368 |
| No | Second booster | Comirnaty | 60-85 years without medical risks | Ref | | 84 | Ref | | 12 |
| No | Second booster | Comirnaty | 60-85 years with medical risks | 0,954 | 0,718 - 1,267 | 42 | 1,186 | 0,657 - 2,140 | 9 |

| | | | | | | | | | |
|---|---|---|---|---|---|---|---|---|---|
| No | Second booster | Spikevax | 60-85 years without medical risks | Ref | | 282 | Ref | | 40 |
| No | Second booster | Spikevax | 60-85 years with medical risks | 1,013 | 0,871 - 1,178 | 173 | 1,091 | 0,712 - 1,669 | 24 |
| Yes | Primary series | Comirnaty | 18-59 years without medical risks | Ref | | 114 | Ref | | 55 |
| Yes | Primary series | Comirnaty | 18-59 years with medical risks | 1,235 | 0,777 - 1,964 | 51 | 1,028 | 0,589 - 1,797 | 28 |
| Yes | Primary series | Comirnaty | 60-85 years without medical risks | 0,866 | 0,571 - 1,311 | 75 | 1,298 | 0,620 - 2,718 | 13 |
| Yes | Primary series | Comirnaty | 60-85 years with medical risks | 1,078 | 0,676 - 1,716 | 52 | 2,986 | 0,973 - 9,161 | 5 |
| Yes | Primary series | Spikevax | 18-59 years without medical risks | Ref | | 37 | Ref | | 74 |
| Yes | Primary series | Spikevax | 18-59 years with medical risks | 0,865 | 0,460 - 1,629 | 14 | 1,701 | 0,919 - 3,146 | 14 |
| Yes | Primary series | Spikevax | 60-85 years without medical risks | 0,084 | 0,029 - 0,243 | 4 | N/A | N/A | 0 |
| Yes | Primary series | Spikevax | 60-85 years with medical risks | 1,639 | 0,559 - 4,802 | 4 | N/A | N/A | 0 |
| Yes | Primary series | Vaxzevria | 18-59 years without medical risks | Ref | | 19 | Ref | | 2 |
| Yes | Primary series | Vaxzevria | 18-59 years with medical risks | 0,781 | 0,226 - 2,699 | 7 | 0,132 | 0,005 - 3,459 | 1 |
| Yes | Primary series | Vaxzevria | 60-85 years without medical risks | 2,206 | 1,054 - 4,609 | 62 | 0,299 | 0,043 - 2,061 | 71 |
| Yes | Primary series | Vaxzevria | 60-85 years with medical risks | 1,362 | 0,621 - 2,989 | 42 | 0,440 | 0,062 - 3,111 | 49 |
| Yes | First booster | Comirnaty | 18-59 years without medical risks | Ref | | 183 | Ref | | 88 |
| Yes | First booster | Comirnaty | 18-59 years with medical risks | 1,126 | 0,882 - 1,438 | 45 | 1,019 | 0,723 - 1,438 | 39 |
| Yes | First booster | Comirnaty | 60-85 years without medical risks | 1,194 | 0,970 - 1,468 | 70 | 1,590 | 1,062 - 2,382 | 25 |
| Yes | First booster | Comirnaty | 60-85 years with medical risks | 1,576 | 1,260 - 1,972 | 58 | 1,536 | 1,021 - 2,312 | 24 |
| Yes | First booster | Spikevax | 18-59 years without medical risks | Ref | | 35 | Ref | | 82 |
| Yes | First booster | Spikevax | 18-59 years with medical risks | 1,169 | 0,834 - 1,637 | 33 | 1,126 | 0,784 - 1,619 | 35 |
| Yes | First booster | Spikevax | 60-85 years without medical risks | 1,084 | 0,839 - 1,401 | 199 | 1,461 | 1,115 - 1,914 | 101 |
| Yes | First booster | Spikevax | 60-85 years with medical risks | 1,186 | 0,908 - 1,553 | 123 | 1,824 | 1,331 - 2,499 | 58 |
| Yes | Second booster | Comirnaty | 60-85 years without medical risks | Ref | | 92 | Ref | | 11 |
| Yes | Second booster | Comirnaty | 60-85 years with medical risks | 1,096 | 0,856 - 1,406 | 76 | 1,331 | 0,617 - 2,872 | 6 |
| Yes | Second booster | Spikevax | 60-85 years without medical risks | Ref | | 221 | Ref | | 31 |
| Yes | Second booster | Spikevax | 60-85 years with medical risks | 0,899 | 0,746 - 1,083 | 142 | 0,814 | 0,502 - 1,320 | 17 |

**Table E. Effect of age and medical risk group on waning.** GMC ratios of interaction with time since vaccination are presented per 30 days. GMC ratio>1 means less waning in that group compared with the reference group; GMC ratio<1 means more waning in that group compared with the reference group.

| Dose | Age and medical risk | GMC ratio Comirnaty | GMC ratio Spikevax | GMC ratio Vaxzevria |
|---|---|---|---|---|
| Primary series | 18-59 years without medical risks | Ref | Ref | Ref |
| | 18-59 years with medical risks | 1.011 (0.963-1.063) | 1.020 (0.966-1.076) | 0.767 (0.590-0.996) |
| | 60-85 years without medical risks | 0.951 (0.913-0.991) | 1.026 (0.855-1.232) | 0.966 (0.857-1.090) |
| | 60-85 years with medical risks | 0.943 (0.899-0.989) | 1.524 (1.191-1.950) | 0.978 (0.866-1.105) |
| First booster | 18-59 years without medical risks | Ref | Ref | |
| | 18-59 years with medical risks | 0.980 (0.920-1.044) | 0.956 (0.895-1.020) | |
| | 60-85 years without medical risks | 1.161 (1.101-1.225) | 1.064 (1.019-1.111) | |
| | 60-85 years with medical risks | 1.195 (1.124-1.272) | 1.088 (1.040-1.139) | |
| Second booster | 60-85 years without medical risks | Ref | Ref | |
| | 60-85 years with medical risks | 0.923 (0.789-1.081) | 0.932 (0.860-1.010) | |

**Table F. Effect of age and immune suppressing conditions on waning.** GMC ratios of interaction with time since vaccination are presented per 30 days. GMC ratio>1 means less waning in that group compared with the reference group; GMC ratio<1 means more waning in that group compared with the reference group.

| Dose | Age and immune compromising conditions | GMC ratio Comirnaty | GMC ratio Spikevax | GMC ratio Vaxzevria |
|---|---|---|---|---|
| Primary series | 18-59 years without immune compromising conditions | Ref | Ref | Ref |
| | 18-59 years with immune compromising conditions | 0.980 (0.858-1.120) | 0.997 (0.889-1.118) | 1.034 (0.132-8.068) |
| | 60-85 years without immune compromising conditions | 0.934 (0.903-0.966) | 1.163 (0.997-1.356) | 1.005 (0.903-1.118) |
| | 60-85 years with immune compromising conditions | 1.033 (0.904-1.180) | 1.042 (0.608-1.786) | 0.984 (0.848-1.141) |
| First booster | 18-59 years without immune compromising conditions | Ref | Ref | |
| | 18-59 years with immune compromising conditions | 0.842 (0.728-0.973) | 0.934 (0.793-1.100) | |
| | 60-85 years without immune compromising conditions | 1.177 (1.127-1.229) | 1.091 (1.053-1.131) | |
| | 60-85 years with immune compromising conditions | 0.896 (0.692-1.162) | 1.024 (0.929-1.129) | |
| Second booster | 60-85 years without immune compromising conditions | Ref | Ref | |
| | 60-85 years with immune compromising conditions | 1.043 (0.747-1.457) | 0.825 (0.693-0.982) | |

**Table G. Effect of prior infection on waning.** GMC ratios of interaction with time since vaccination are presented per 30 days. GMC ratio>1 means less waning in that group compared with the reference group; GMC ratio<1 means more waning in that group compared with the reference group.

| Dose | Prior infection | GMC ratio Comirnaty | GMC ratio Spikevax | GMC ratio Vaxzevria |
|---|---|---|---|---|
| Primary series | No | Ref | Ref | Ref |
| | Yes | 1.018 (0.971-1.068) | 0.836 (0.781-0.896) | 1.152 (1.079-1.231) |
| First booster | No | Ref | Ref | |
| | Yes | 1.130 (1.082-1.179) | 1.008 (0.972-1.047) | |
| Second booster | No | Ref | Ref | |
| | Yes | 1.117 (0.995-1.254) | 0.994 (0.933-1.059) | |
